# Vagus nerve cross-sectional area in patients with Parkinson’s Disease – an ultrasound case-control study

**DOI:** 10.1101/2021.02.15.21251731

**Authors:** Jacob Horsager, Uwe Walter, Tatyana D Fedorova, Katrine B. Andersen, Casper Skjærbæk, Karoline Knudsen, Niels Okkels, Paul von Weitzel-Mudersbach, Stig Eric Dyrskog, Bo Bergholt, Per Borghammer

## Abstract

**Background:** Vagal parasympathetic neurons are prone to degeneration in Parkinson’s disease (PD). High-resolution ultrasound can precisely estimate the cross-sectional area (CSA) of peripheral nerves. Here, we tested the hypothesis that vagus CSA is reduced in PD.

**Methods:** We included 56 healthy controls (HCs) and 63 patients with PD. Using a high-end ultrasound system equipped with a high-frequency transducer, five images were obtained of each nerve. The hypoechoic neuronal tissue was delineated offline with dedicated software and the CSA extracted.

**Results:** In the initial PD vs. HC comparison, no statistically significant differences were observed in mean left vagus CSA (HC: 1.97mm^2^, PD: 1.89mm^2^, P=0.36) or in mean right vagus CSA (HC: 2.37mm^2^, PD: 2.23mm^2^, P=0.17). The right vagus CSA was significantly larger than the left vagus CSA in both groups (P<0.0001). Females were overrepresented in the HC group and presented with generally smaller vagus CSAs. Consequently, sex-adjusted CSA was significantly smaller for the right vagus nerve of the PD group (P=0.041), but not for the left.

**Conclusion:** A small but significant reduction in sex-adjusted right vagus CSA was observed in patients with PD. The left vagus CSA was not significantly reduced in patients with PD. Ultrasound may not be a suitable method to detecting vagal axonal loss in individual patients.

## 1. Introduction

The vagus nerve may play a crucial role in the pathogenesis of Parkinson’s disease (PD). It has been hypothesized that α-synuclein aggregates form in the enteric nervous system, and spread via the autonomic nervous system to the CNS [1]. In this context, vagal parasympathetic neurons that project to most of the gastrointestinal tract serve as entrance to the dorsal motor nucleus of the vagus (DMV). In support, truncal vagotomy decrease the risk of developing PD [2, 3]. The DMV and the vagus nerve itself are among the most frequently affected structures in post-mortem PD studies [4-7]. This is accompanied by up to 50% DMV motor neuron loss in deceased patients [8-10].

Using ^11^C-donepezil PET/CT, we have previously demonstrated decreased gastrointestinal acetylcholinesterase density in patients with PD [11-13]. This is believed to reflect parasympathetic denervation, mainly derived from the vagus nerve. Furthermore, subjects with isolated REM-sleep behavior disorder, the strongest marker of prodromal PD [14], also exhibit decreased gastrointestinal acetylcholinesterase density [15]. Hence, the vagal parasympathetic system is affected years before diagnosis in some patients. Therefore, precise evaluation of vagal degeneration could serve as tool to diagnose PD patients, even during the pre-motor phase.

High-resolution ultrasound is a non-invasive, radiation-free alternative to assess vagal degeneration. The method can precisely measure the caliber of superficial nerves. Six previous studies tested the hypothesis that vagal neuron loss is detectable by ultrasound, but reported conflicting results. Three studies found significant atrophy of both left and right vagus nerves in patients with PD [16-18]. In contrast, three studies reported no difference between patients with PD and healthy controls (HCs) [19-21]. Thus, whether ultrasound can detect vagal neuronal loss in patients with PD remains unresolved.

In the present study we determined the vagus cross-sectional area (CSA) in the largest cohort of PD patients studied so far and compared to HCs. Five images of each nerve were obtained to optimize accuracy and all analyses were performed offline for optimal precision. We compared the vagus CSA with measures of gastrointestinal acetylcholinesterase density (parasympathetic innervation) and retained radiopaque markers (estimate of gastrointestinal transit time) in subgroups. These measures should theoretically be altered in parallel to vagal parasympathetic depletion.

## 2. Methods

### 2.1 Study population

We included 63 early-to-moderate stage PD patients and 56 HCs. Participants were enrolled between April 2019 and November 2020 from ongoing studies and one published study [13] at Aarhus University Hospital, Denmark. Demographic and clinical information is listed in **Table 1**. All participants provided written informed consent. The study was approved by the Science Ethical Committees of the Central Denmark Region.

**Table 1.** Demographic and clinical information.

|  | HC |  | PD |  | P-value |
| --- | --- | --- | --- | --- | --- |
| Sample size *n* | 56 |  | 63 |  |  |
| Sex *m/f* | 26/30 |  | 43/20 |  | **0.025** |
| Age | 69.1 (8.9) |  | 67.4 (7.6) |  | 0.25 |
| Time since diagnosis [months] | - |  | 8 (2-85) |  |  |
| UPDRS III | - |  | 23 (10) |  |  |
| H&Y I/II/III | - |  | 14/43/6 |  |  |
| LEDD [mg] |  |  | 656 (404) |  |  |
| BMI | 24.9 (3.7) |  | 26.0 (4.1) |  | 0.14 |
| Left vagus CSA [mm<sup>2</sup>] | 1.97 (0.48) |  | 1.89 (0.48) |  | 0.36 |
| Right vagus CSA [mm<sup>2</sup>] | 2.37 (0.57) |  | 2.23 (0.55) |  | 0.17 |
| Vagus CSA-symmetry index | 1.17 (1.09-1.39) |  | 1.19 (1.01-1.37) |  | 0.69 |
|  | *Females* | *Males* | *Females* | *Males* |  |
| Left vagus CSA [mm<sup>2</sup>] | 1.86 (0.42) | 2.09 (0.52) | 1.81 (0.43) | 1.92 (0.50) | - |
| Right vagus CSA [mm<sup>2</sup>] | 2.23 (0.47) | 2.54 (0.63) | 2.01 (0.42) | 2.33 (0.58) | - |

H&Y = Hoehn and Yahr stage, LEDD = Levodopa equivalent daily dose, BMI = Body Mass Index, CSA = cross-sectional area. Vagus CSA-symmetry index were calculated as right/left. Results are presented as mean (SD) or median (IQR).

### 2.2 Vagus nerve ultrasound

All images were obtained with a high-end ultrasound system (Philips EPIQ 7, *Andover, MA, USA*) equipped with a linear array 12.0-MHz transducer (L12-3, Phillips, *Bothell, VA, USA*). Participants were examined in the supine position by the same investigator (JH), who was not blinded to clinical status at ultrasound examination. The vagus nerve was identified mid-cervically at the level of the thyroid cartilage. On approximately the same location (within a few centimeters), five individual images were obtained bilaterally, i.e. ten images of each participant. All images were exported to offline analysis. In PMOD 4.0 (*PMOD, Zürich, Switzerland*), the color scale of each image was adjusted to the maximum intensity of the vagus nerve rim. This was performed to ensure optimal contrast between hypoechoic neuronal tissue and the hyperechoic epineurium. Next, the inner border of the hyperechoic epineurium was delineated and CSA was extracted in mm^2^ with two decimals (**Figure 1**). Of the five images of each nerve, the largest and smallest CSA were discarded to prevent influence of potential outliers and the average of the three middle values was used for statistical analyses. All offline image analyses were performed blinded to clinical status.

**Figure 1.**
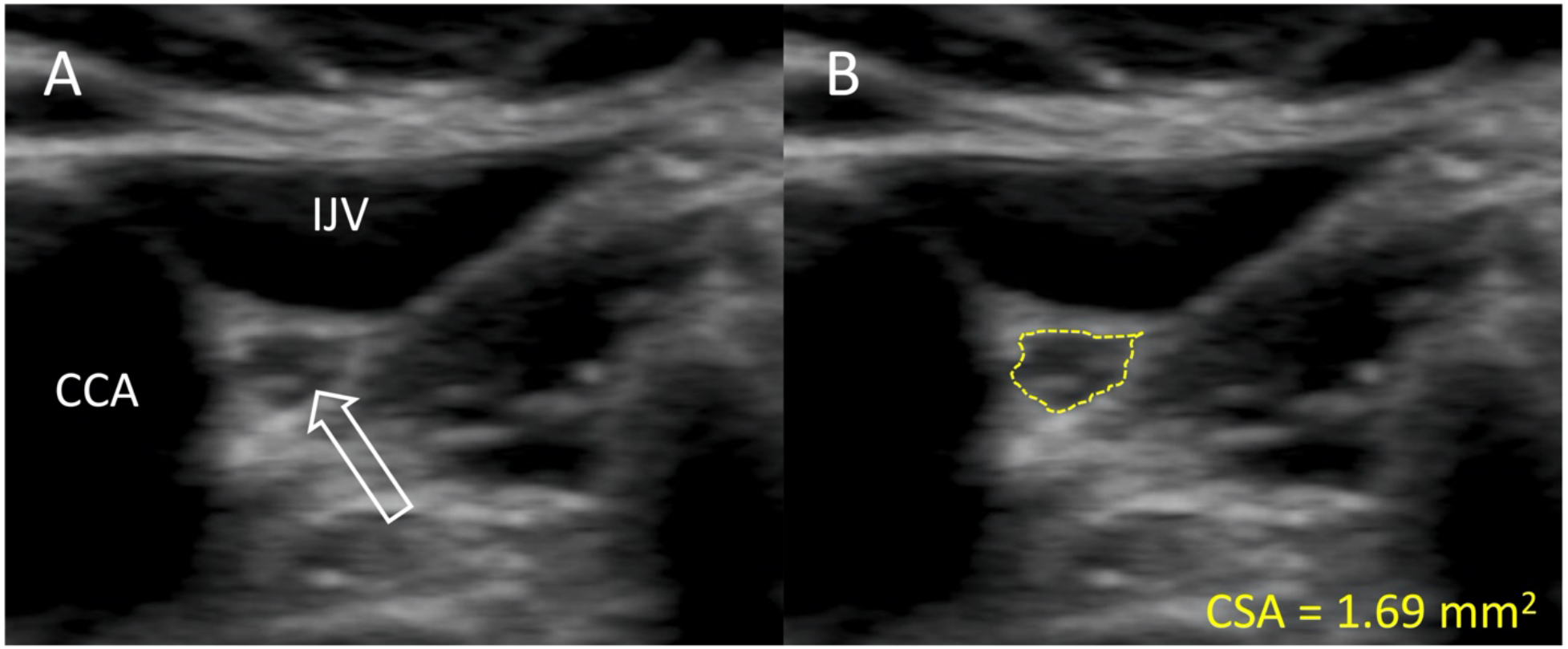
Representative ultrasound image of the vagus nerve. **A:** The vagus nerve (white arrow) of a patient with PD. **B:** Delineation of the same vagus nerve following the hyperechoic epineurium. IJV = internal jugular vein, CCA = common carotid artery, CSA = cross-sectional area.

### 2.3 Other measurements

Radiopaque marker (ROM) data was available for 74 subjects (PD=53, HC=21) from two on-going studies and one published study [13]. In short, ten ROMs were ingested every morning, six days prior to an abdominal CT scan. The number of retained ROMs estimates the colonic transit time. Colonic acetylcholinesterase density (measure of parasympathetic innervation) assessed by ^11^C-donepezil PET/CT, was available in 41 patients with PD [13]. These objective measures of gastrointestinal function and innervation was used to correlate with vagus CSA in subgroup analyses.

### 2.4 Statistical analyses

Statistical analyses were performed with Stata 13.1 and GraphPad Prism 7.0. Normality in distribution was assessed by Q-Q plots and histograms. Group differences of sex were investigated using Fisher’s exact tests. Group differences of age and BMI were tested with student’s *t-*test. Differences between right and left side were assessed with paired *t-*tests. Differences in symmetry index were assessed with Mann-Whitney-U test. Pearson correlation coefficients were calculated to explore correlations between vagus CSA and other variables (age, BMI, time since diagnosis, disease severity defined by Hoehn and Yahr stage (H&Y), ROM, and colon acetylcholinesterase density measured by ^11^C-donepezil PET). The unmatched composition of our two cohorts regarding sex was addressed with a multiple linear regression model to calculate sex-adjusted differences in CSA. This model was checked by diagnostic plots of residuals. P-values <0.05 were considered significant. Method precision was ascertained with coefficient of variation (SD/mean) for each nerve using the three middle CSA values.

## 3. Results

Mean vagus nerve CSAs are presented in **Table 1**, and depicted graphically in **Figure 2**. There was no significant difference in the crude comparison between patients with PD and HCs. We observed a tendency towards higher CSA in males than females, most pronounced in the right vagus (**Table 1**). Thus, we used multiple linear regression to determine sex-adjusted CSA of the vagus. First, we applied a model allowing different slopes in each group. We found no significant slope interactions (P>0.5). Thus, we applied a model with one common slope. Sex-adjusted difference in mean left vagus CSA was 0.12mm^2^ (95%CI: −0.06 – 0.29mm^2^, P=0.19) and right vagus was 0.21mm^2^ (95%CI: 0.01 – 0.41mm^2^, P=0.041). In this model, female sex was a significant negative predictor of right vagus CSA (P=0.003), but not of the left vagus CSA (P=0.06). No difference in symmetry index was observed (P=0.69).

**Figure 2.**
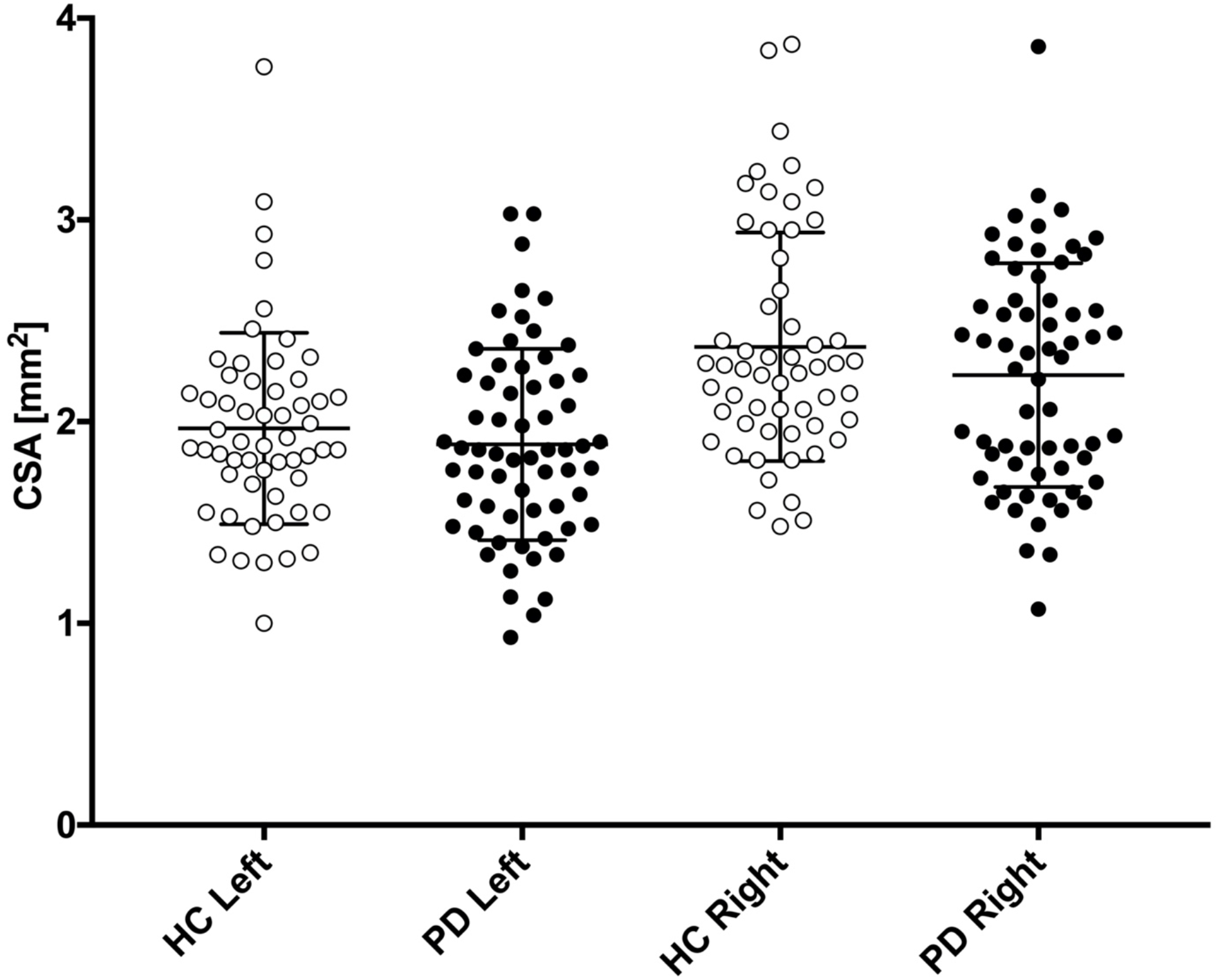
Left and right vagus nerve cross-sectional area (CSA) is presented for patients with Parkinson’s disease (PD, black circles) and healthy controls (HC, white circles). Mean and standard deviation is depicted with black error bars.

In the HC group, mean right vagus CSA was 0.4mm^2^ larger than mean left vagus CSA (95%CI: 0.24 – 0.57mm^2^, P<0.0001). In the PD group, mean right vagus CSA was 0.34mm^2^ larger than left vagus CSA (95%CI: 0.20 – 0.48mm^2^, P<0.0001). Left- and right vagus CSA did not correlate with age, BMI, time since diagnosis, disease severity (H&Y), ROM (estimate of colonic transit time), or colon acetylcholinesterase density measured by ^11^C-donepezil PET (P>0.05). Mean coefficient of variation of vagus CSAs was 6.7%.

## 4. Discussion

In the present study we found no difference in vagus nerve CSA between patients with PD and HCs, but female vagus CSA was smaller than male vagus CSA in both groups. One previous report found the opposite pattern; vagus CSA in females were larger than males [22]. More females were included in our HC group compared to the PD group. Hence, we determined the sex-adjusted differences between patients with PD and HCs, and found a statistically significant reduction of the right vagus CSA (P=0.041) but not of the left vagus CSA (P=0.19).

Although the mean right vagus CSA was significantly reduced in PD, the effects size of ∼ 10% is small. The effect size in other studies, which reported statistically significant CSA reduction, range from approximately 10-30% [16-18]. Neuronal loss in DMV has been reported to be up to 50% in post-mortem studies [8-10]. Other nuclei related to the vagus nerve (nucleus ambiguus, solitary nucleus, and spinal trigeminal nucleus) are less affected by α-synuclein aggregates [23]. Also, no motor neurons are lost in nucleus ambiguus [9, 24]. The axonal diameter of somato-motor and somato-sensory myelinated fibers is generally much larger than that of autonomic neurons, and must necessarily occupy a substantial fraction of the mid-cervical vagus CSA. Cardiac sympathetic neurons also reside in the vagus nerve and occupy up to 5% of the CSA [25, 26]. Cardiac sympathetic neurons are particularly vulnerable in PD and may also contribute to slight vagal atrophy [27]. However, if vagal atrophy is mainly caused by sympathetic and parasympathetic (DMV) axonal loss, the reduction in CSA is probably very small, and could explain why three studies failed to show vagal atrophy in PD [19-21]. Additionally, the vagus nerve shows frequent branching (up to 29%) [28]. Branched nerves have smaller CSA at the mid-cervical level [28]. Although we have no reasons to believe that branching should differ between patients with PD and HC, it may have contributed to increased data variance.

Time since diagnosis could be another factor explaining some incongruence between studies. In the present study the median duration was short (8 months), as many of the patients were newly diagnosed. The three previous studies that demonstrated significant CSA reduction included patients 5-10 years post diagnosis [16-18]. Thus, the rather small difference demonstrated in the present study might be partly caused by the shorter duration of motor symptoms. However, we found no correlation between vagus CSA and time since diagnosis in the present data, and the three studies that failed to show a significant difference also investigated moderately affected patients, 4-8 years post diagnosis [19-21].

If a biological difference in right vagus CSA truly exists between patients with PD and HCs, two aspects may have enabled us to identify this small difference in the present study; 1) our 2-3 times larger PD cohort than previous studies, and 2) offline measurements as advised for optimal precision [29]. However, our study does not support that this method can be applied for meaningful interpretation at the individual level, because the average between-group difference of 0.21mm^2^ is hardly discerned, given that the normal vagal CSA ranges from 1mm^2^ to nearly 4 mm^2^ (**Figure 2**).

Right vagus CSA was significantly larger than left vagus CSA as previously reported [16-19, 21, 22, 29]. This also corresponds to the side-to-side difference of the cervical vagus neuronal area defined on post-mortem [30]. Further, the right mid-cervical vagus contains ∼20% more nerve fibers than the left [31]. The mean CSA values were in reasonable agreement with six previous studies that estimated vagus CSA by tracing the epineural rim, as in the present study [16, 18, 19, 21, 22, 29]. Thus, in studies using the tracing method, a reasonable inter-study reproducibility of the vagus CSA seems to exists.

Vagus CSA did not correlate with the number of retained ROM, nor with colon acetylcholinesterase density measured by ^11^C-donepezil PET. If vagus CSA atrophy occurs in parallel to parasympathetic denervation, we would expect these parameters to be correlated. However, not all participants had ROM and ^11^C-donepezil PET data available which could explain the absence of correlation between these parameters.

The precision of the three middle values was evaluated for each nerve by calculating the coefficient of variation. The average coefficient of variation was acceptable (6.7%). We also calculated the average distance from the three middle values to the mean. The average distance was 4.9% of the mean, e.g. values defining a CSA of 2mm^2^ on average were 0.1mm^2^ from the “exact” value. Therefore, we argue that obtaining five ultrasound images of each nerve and using the three middle values to determine the CSA yields precise results.

This study has some limitations. First, the ultrasound investigator was not blinded to clinical status during examination. However, the offline CSA estimation was performed blinded. Second, our two groups were matched on age but not sex, forcing us to do linear adjustment in the comparison analyses.

In conclusion, we found a small but significant reduction in sex-adjusted right vagus CSA in patients with PD. No difference was found in left vagus CSA. Sex was a significant predictor for the right-but not left vagus caliber. This study does not support the use of vagus ultrasound as a diagnostic marker for PD, but there may be applications for the method in a research setting.

## Data Availability

Data will be available upon request to the corresponding author.

